# Factors associated with awareness and acceptability of pre-exposure prophylaxis among women of reproductive age in Kenya: an analysis of the 2022 Kenya demographic and health survey

**DOI:** 10.1101/2025.08.01.25332793

**Authors:** Lilian Nuwabaine, John Baptist Asiimwe, Earnest Amwiine, Robert Kiiza, Angella Namulema, Imelda Namatovu, Joseph Kawuki

## Abstract

**Background:** Human Immunodeficiency Virus (HIV) remains a significant public health challenge in Kenya, with women of reproductive age (WRA) at an increased risk. Although pre-exposure prophylaxis (PrEP) was integrated into Kenya’s HIV prevention program close to a decade ago, it is still underutilized among WRA outside the key populations at a high risk for HIV. Understanding PrEP awareness and acceptability is crucial for scaling prevention efforts. This study, therefore aimed to determine the factors associated with PrEP awareness and acceptability among WRA using the 2022 Kenya Demographic and Health Survey (KDHS).

**Methods:** Secondary data comprising 16,638 WRA from the 2022 KDHS, which used a two-stage stratified sampling design was analyzed using univariable and multivariable logistic regression models in SPSS (version 29).

**Results:** 48.4% (95% CI: 47.2–49.7) of WRA were aware of PrEP, and 75.0% (95% CI: 73.3–76.6) approved its use. Higher education (tertiary: Adjusted odds ratio (AOR): 2.11, 95% CI: 1.50–2.97), good HIV knowledge (AOR 2.70, 95% CI: 1.67–4.36), living with HIV (AOR 3.79, 95% CI: 1.59–9.03) and multiple sexual partners (AOR 2.29, 95% CI: 1.44–3.64) were associated with high odds of PrEP awareness compared with their counterparts. Women from Nyanza (AOR 2.61, 95% CI: 1.61–4.22) and Northeastern regions, and those from the Luo tribe (AOR 2.51, 95% CI: 1.57–4.02), also had high odds of awareness than those from the Coastal region and Kikuyu tribe, respectively. Muslim women (AOR 0.51, 95% CI: 0.31–0.84) and those who justified wife-beating (AOR 0.63, 95% CI: 0.51–0.76) were less likely to be aware of PrEP. Compared with their counterparts, we found high odds of PrEP acceptability among women who had their first sexual intercourse at ≥25 years (AOR 40.06, 95% CI: 14.59–109.99), had good HIV knowledge (AOR 4.88, 95% CI: 1.28–18.62) and lived in Nyanza province (AOR 8.76, 95% CI: 3.27–23.46).

**Conclusion:** We found a low PrEP awareness level among WRA when compared with its acceptability. Sociodemographic, behavioral, and psychosocial factors seem to influence PrEP awareness and acceptability. Targeted interventions, including integrating PrEP education in healthcare, school sexuality curriculum, workplaces, regional and culturally specific mass media campaigns, and addressing socio-cultural barriers, are vital to enhance uptake and reduce HIV incidence.

## Background

HIV remains a critical public health challenge, particularly in Sub-Saharan Africa, which accounts for over two-thirds of the global HIV burden, with women comprising 53% of the 39 million people living with HIV worldwide (1). Kenya, with the seventh-largest HIV burden globally, had 1.3 million people living with HIV/AIDS, with women accounting for 67% of the 22,154 new infections reported in 2022 (1). Adolescent girls and young women are disproportionately affected, facing a fivefold higher likelihood of HIV infection compared with men, driven by gender inequality, economic dependence, intimate partner violence, transactional relationships, and low condom use (2).

Pre-exposure prophylaxis (PrEP) is a highly effective HIV prevention strategy for populations at substantial HIV risk (3). Available in oral, injectable, and vaginal rings, PrEP reduces HIV acquisition risk by up to 75% when used consistently, as demonstrated in clinical trials (4). Since its introduction in Kenya in 2016 and integration into the national HIV prevention program in 2017, PrEP has been prioritized for key and priority populations at high risk for HIV, including adolescent girls and young women (AGYW), female sex workers (FSWs), and serodiscordant couples unlike other women in the general population (2, 5).

Despite progress in PrEP scale-up, with approximately 44,000 Kenyans using PrEP by 2020, awareness and uptake among women remain low, particularly among WRA not classified as key and priority populations at high risk for HIV (6, 7). Studies indicate that only 34.29% of Kenyan women are aware of PrEP, with uptake rates as low as 4% among eligible AGYW and 27% among serodiscordant couples (8, 9). Barriers to PrEP awareness, acceptability, and use included low HIV risk perception, concerns about pill burden, side effects, and intimate partner violence, while facilitators included PrEP education, healthcare provider support, and social networks (8, 10). In contrast, PrEP awareness and acceptability also varied across SSA, with Tanzania reporting 8% awareness among women and Nigeria 7.8% among women in the general population (11, 12). These inconsistencies highlight the need for population-based data to inform targeted interventions.

The limited focus on PrEP awareness and acceptability among the general population of WRA in Kenya represents a critical research gap. Most studies targeted key and priority populations at high risk for HIV attending specialized clinics, with the concept remaining under-explored in the general population of WRA (13, 14). Understanding the factors influencing PrEP awareness and acceptability among WRA is essential for reducing HIV stigma, increasing PrEP uptake, and scaling up prevention efforts in general. Consequently, this is vital for reducing HIV incidence among women, in line with the global goal of reducing new infections to fewer than 370,000 annually by 2025 (1). The 2022 KDHS provides a nationally representative dataset to investigate these determinants, offering insights into factors influencing PrEP awareness and acceptability among WRA. Therefore, this study aimed to address this gap by investigating the sociodemographic, lifestyle behavioral, sexual behavioral, psychosocial, and intimate partner violence factors associated with PrEP awareness and acceptability among WRA in Kenya. The study findings may contribute to evidence-based strategies for HIV prevention in Kenya and other sub-Saharan African countries.

## Methods

### Data source, sample design, and collection

We analysed the 2022 Kenya Demographic and Health Survey (KDHS) data, which used a two-stage stratified sampling approach. In the first stage, 1692 enumeration areas (EAs) or clusters were chosen from a comprehensive sample frame of 129,067 EAs based on the 2019 Kenya population and housing census, utilizing equal probability with independent selection(15). This was succeeded by compiling a list of houses to establish a sampling frame that was utilized in the next phase to choose 25 households from each cluster. However, all households in a cluster were sampled if they were less than 25. In total, the survey was implemented in 1691 clusters. The Inner-City Fund (ICF) facilitated the pretesting of the study tools and the training of data collectors, and data was collected between February and July 2022. All WRA between the ages of 15 and 49 who were regular members of the chosen households, or those who had spent the previous night in those households, were interviewed in either English or Swahili (15). Out of 33,137 eligible women, 16,638 women responded to the survey and were included in this analysis (50.2 % response rate) (15). Even though the dataset included numerous variables, we focused solely on those that were pertinent to our research.

### Study variables

#### Dependent/outcome variables

The two primary outcomes of this study were PrEP awareness and acceptability as captured by the following survey questions: 1) Have you heard of PrEP, a medicine taken daily that can prevent a person from getting HIV? and 2) Do you approve of people who take a pill every day to prevent getting HIV? Women who answered yes were coded as 1; otherwise as 0.

#### Independent variables

The factors included in the analysis were categorized into five, namely, sociodemographic, lifestyle behavioral, sexual behavioral, psychosocial, and intimate partner violence factors. The 15 sociodemographic factors that were included in the analysis, are; age in years (15–24, 25–34, 35–59), wealth index (poorest, poorer, middle, richer, and richest), education (none/primary, secondary or tertiary), working status (Yes vs No), residence (rural vs urban), religion (Christian, Muslim, or others), marital status (married/cohabiting, or unmarried), region (categorized into Kenya’s eight provinces; Northeastern, Coast, Eastern, Rift Valley, Western, Central, Nairobi and Nyanza, ), household size (≥5 vs ≤4 members), and ethnicity (categorized into Kenya’s 12 tribes; Kalenjin, Embu, Kamba, Kikuyu, Luhya, Kisii, Maasai, Luo, Mijikenda/Swahili, Meru, Taita/Taveta, Somali, and Others). Using principal component analysis, the wealth index was calculated by 2022 KDHS from information on household asset ownership (15). Additionally, the husbands ‘education and working status were also included in the analysis.

Autonomy was assessed through four proxy variables namely, who heads the household (male vs female), who makes decisions related to healthcare-seeking, major household purchases, and earnings (jointly with partner or another person, self, partner, or others). The participants’ perceived health status at the time of the interview (good, moderate, bad), and exposure to mass media such as television, radio, newspapers, and internet (No vs Yes) were also included in the analysis.

Three lifestyle behavioral factors were also analysed: whether the participants and their sex partners (husbands) consumed alcohol or used tobacco (yes/no). We also examined nine variables that were related to sexual behaviors which included the age of the participant at first sexual intercourse in years (≤14, 15-24, ≥25), recent sexual activity (active vs inactive), current pregnancy (yes/no), number of sex partners in last 12 months (1 partner vs ≥2 partners), number of living children (≤2, 3-4, ≥5), number of lifetime sex partners (≤5, 6-10, >10), HIV serostatus (negative vs positive), and whether one had gotten any symptoms of sexually transmitted infections (STIs) in the last 12 months preceding the survey in the form of a genital ulcer or discharge (yes/no). Additionally, this study sought to investigate if the use of other HIV prevention services such as condom use at last sex (yes/no), and HIV testing and counseling (yes/no) had an influence on PrEP awareness and acceptability.

Four psychosocial factors were included in the analysis such as the desire for more children (wants, does not want, undecided), whether their wives were justified to refuse sex if they had other women (yes/no), HIV knowledge, and stigma. Knowledge of HIV as a binary variable was computed from 5 items about HIV transmission and prevention with answer options of “Yes”, “No” or “Don’t Know”. A correct response for each item was awarded a score of 1 with a maximum score of 5 which was further categorized as poor (≤2), or good knowledge (≥3). Whereas HIV stigma was operationalized as having stigmatizing attitudes towards people living with HIV, measured by the item “Would you buy vegetables from a vendor with HIV” (yes/no).

We sought to establish if intimate partner violence (IPV) related factors had an influence on PrEP awareness and acceptability among women of reproductive age by including in the analysis, variables such as the participant being afraid of their husband/partner (yes/no), justified beating (yes/no), and experiencing economic, controlling, emotional, physical and sexual forms of IPV (yes/no). It is worth noting that justified beating (5 items), controlling (5 items), emotional (4 items), physical (7 items), and sexual (3 items) IPV variables were composite variables constructed from numerous binary response items, where an agreement to any of the statements or items (≥1) indicated justified beating or an experience of any forms of IPV.

### Statistical analysis

The complex samples feature in SPSS (V29) was utilized to analyze the data, considering the intricate sample design characteristic of DHS data. (16, 17). The complex sample package yields accurate parameter estimates by considering the sample stratification, clustering, and weighting that were employed when selecting the study participants (16, 17). Furthermore, to address the unequal probability of sampling across different strata and to guarantee that the study results are representative, DHS sample weights were utilized for all computed frequencies (16, 17). Prior to conducting the analysis, the data underwent a cleaning process and the creation of dummy variables. Descriptive statistics, including frequencies, were calculated for all categorical variables at the univariate level. Both univariate and multivariate logistic regression analyses were performed to identify independent factors related to PrEP awareness and acceptability. All variables that had P-values less than 0.05 were included in a simple multivariate logistic regression to identify the factors linked to the two outcome variables. The odds ratios for all variables are presented along with their 95% confidence intervals. Additionally, multi-collinearity among all predictor variables in the model was evaluated using a variance inflation factor (VIF), with a threshold of greater than 10 as the cutoff(16, 17). None of the factors surpassed the threshold.

### Ethical consideration

No ethical approval was required to examine the secondary data since it is publicly accessible. Nevertheless, authorization to access the 2022 KDHS datasets was granted by MEASURE DHS (https://www.dhsprogram.com/data/available-datasets.cfm). Approval for the study detailed in the datasets was granted by the Institutional Review Board of the Inner-City Fund (ICF). The Kenya National Bureau of Statistics carried out the study in partnership with various stakeholders.

## Results

### Socio-demographic Characteristics of the study participants

In total, 16638 women of reproductive age were included in this analysis (**Table 1**). Most of the participants were aged 15-34 years (68.6%), lived in rural settings (58.9%), and identified as being from the Rift Valley, Eastern, Central, and Nairobi provinces (66.4%). The majority identified as being from the Kikuyu, Luhya, Kalenjin, and Kamba tribes (62.8%), were Christians by faith (88.9%), married (55.8%), working (52.8%), and had completed utmost secondary education (80.8%). In total, 66.8% belonged to the middle, richer, and richest quintiles, 54.5% had more than five household members, and 60.8% lived in male-headed households. Most participants made decisions to seek healthcare services (43.9%), make household purchases (59.2%), and spend earnings (51.2%) jointly with their partners. The majority were exposed to mass media, which included radio (75.6%), television (66.3%), the internet (47.6%), and newspapers (19.6%), and described themselves as being in good health status (78.0%). In total, 92.1% of the participating women had husbands who were working and had utmost attained secondary education (76.3%).

**Table 1.** Socio-demographic characteristics of the study participants.

| **Variable** | ***n*** | ***Weighted %*** |
| --- | --- | --- |
| **Sociodemographic factors** |  |  |
| **Age (years)** |  |  |
| 15-24 | 6140 | 36.9 |
| 25-34 | 5269 | 31.7 |
| 35-59 | 5230 | 31.4 |
| **Residence** |  |  |
| Rural | 9801 | 58.9 |
| Urban | 6837 | 41.1 |
| **Region/Province** |  |  |
| Coast | 1491 | 9.0 |
| Northeastern | 359 | 2.2 |
| Eastern | 2199 | 13.2 |
| Central | 2180 | 13.1 |
| Rift Valley | 4504 | 27.1 |
| Western | 1755 | 10.5 |
| Nyanza | 1993 | 12.0 |
| Nairobi | 2157 | 13.0 |
| **Ethnicity** |  |  |
| Embu | 194 | 1.2 |
| Kalenjin | 2820 | 17.0 |
| Kamba | 1913 | 11.5 |
| Kikuyu | 2990 | 18.0 |
| Kisii | 1001 | 6.0 |
| Luhya | 2711 | 16.3 |
| Luo | 2187 | 13.2 |
| Maasai | 383 | 2.3 |
| Meru | 987 | 5.9 |
| Mijikenda/Swahili | 848 | 5.1 |
| Somali | 453 | 2.7 |
| Taita/Taveta | 122 | 0.7 |
| Other | 6 | 0.0 |
| **Education** |  |  |
| None/primary | 6967 | 41.9 |
| Secondary | 6464 | 38.9 |
| Tertiary | 3207 | 19.3 |
| **Religion** |  |  |
| Christians | 14788 | 88.9 |
| Muslims | 1183 | 7.1 |
| Others | 667 | 4.0 |
| **Marital status** |  |  |
| Unmarried (single divorced/widowed/separated) | 7354 | 44.2 |
| Married/Cohabiting | 9284 | 55.8 |
| **Working status/occupation** |  |  |
| Not working | 7851 | 47.2 |
| Working | 8788 | 52.8 |
| **Wealth index** |  |  |
| Poorest | 2568 | 15.4 |
| Poorer | 2956 | 17.8 |
| Middle | 3069 | 18.4 |
| Richer | 3722 | 22.4 |
| Richest | 4323 | 26.0 |
| **Sex of Household head** |  |  |
| Male | 10114 | 60.8 |
| Female | 6524 | 39.2 |
| **Household number** |  |  |
| ≤4 | 7572 | 45.5 |
| ≥5 | 9066 | 54.5 |
| **Health status** |  |  |
| Bad | 425 | 2.6 |
| Moderate | 3234 | 19.4 |
| Good | 12979 | 78.0 |
| **Health seeking decision making** |  |  |
| Self | 3994 | 43.0 |
| Joint | 4073 | 43.9 |
| Partner | 1197 | 12.9 |
| Others | 20 | 0.2 |
| **Purchases decision making** |  |  |
| Self | 1867 | 20.1 |
| Joint | 5499 | 59.2 |
| Partner | 1882 | 20.3 |
| Others | 37 | 0.4 |
| **Earnings decision making** |  |  |
| Self | 634 | 7.2 |
| Joint | 4510 | 51.2 |
| Partner | 3660 | 41.6 |
| Other | 4 | 0.0 |
| **Husband Education** |  |  |
| None/primary | 4027 | 43.8 |
| Secondary | 2984 | 32.5 |
| Tertiary | 2180 | 23.7 |
| **Husband's Working status/occupation** |  |  |
| Not working | 708 | 7.9 |
| Working | 8282 | 92.1 |
| **Newspaper** |  |  |
| No | 13378 | 80.4 |
| Yes | 3260 | 19.6 |
| **TV** |  |  |
| No | 5613 | 33.7 |
| Yes | 11025 | 66.3 |
| **Radio** |  |  |
| No | 4052 | 24.4 |
| Yes | 12586 | 75.6 |
| **Internet use** |  |  |
| No | 8715 | 52.4 |
| Yes | 7923 | 47.6 |
| **Lifestyle Behavioural factors** |  |  |
| **Tobacco use** |  |  |
| No | 16467 | 99.0 |
| Yes | 171 | 1.0 |
| **Alcohol use** |  |  |
| No | 15733 | 99.7 |
| Yes | 43 | 0.3 |
| **Husband drinks Alcohol** |  |  |
| No | 3511 | 69.4 |
| Yes | 1548 | 30.6 |
| **Sexual Behavioural factors** |  |  |
| **Age at first sex (years)** |  |  |
| ≤14 | 2063 | 14.6 |
| 15-24 | 11658 | 82.7 |
| ≥25 | 370 | 2.6 |
| **Sexual activity** |  |  |
| Inactive | 8606 | 51.7 |
| Active | 8031 | 48.3 |
| **Number of sex partners in last 12 months** |  |  |
| 1 partner | 11642 | 95.2 |
| ≥2 partners | 583 | 4.8 |
| **Number of lifetime sex partners** |  |  |
| <5 | 12780 | 90.7 |
| 05-Oct | 1025 | 7.3 |
| >10 | 290 | 2.1 |
| **HIV status** |  |  |
| Negative | 13833 | 97.7 |
| Positive | 332 | 2.3 |
| **Genital ulcer in last 12 months** |  |  |
| No | 15884 | 95.6 |
| Yes | 738 | 4.4 |
| **Genital Discharge in the last 12 months** |  |  |
| No | 15267 | 91.9 |
| Yes | 1347 | 8.1 |
| **Number of living children** |  |  |
| ≤2 | 10652 | 64.0 |
| 03-Apr | 3916 | 23.5 |
| ≥5 | 2071 | 12.4 |
| **Use of other HIV services** |  |  |
| **Condom use at Last Sex** |  |  |
| No | 12687 | 90.0 |
| Yes | 1408 | 10.0 |
| **Ever tested for HIV** |  |  |
| No | 2355 | 14.2 |
| Yes | 14283 | 85.8 |
| **Pregnant** |  |  |
| No or unsure | 15712 | 94.4 |
| Yes I am | 926 | 5.6 |
| **Psychosocial factors** |  |  |
| **Desire for children** |  |  |
| Wants | 9326 | 57.8 |
| Does not want | 5789 | 35.9 |
| Undecided | 1029 | 6.4 |
| **HIV knowledge** |  |  |
| Poor | 459 | 4.0 |
| Good | 10942 | 96.0 |
| **Would buy vegetables from vendors with HIV** |  |  |
| Yes (No stigma) | 3682 | 22.6 |
| No (stigma) | 12639 | 77.4 |
| **Wife justified in asking her husband to use a condom** |  |  |
| No | 2439 | 15.0 |
| Yes | 13870 | 85.0 |
| **IPV factors** |  |  |
| **Respondent afraid of Husband** |  |  |
| No | 3615 | 22.2 |
| Yes | 1444 | 8.9 |
| **Justified beating** |  |  |
| No | 11282 | 67.9 |
| Yes | 5324 | 32.1 |
| **IPV forms** |  |  |
| **Emotional IPV** |  |  |
| No | 3582 | 70.8 |
| Yes | 1477 | 29.2 |
| **Controlling IPV** |  |  |
| No | 1963 | 38.8 |
| Yes | 3096 | 61.2 |
| **Physical IPV** |  |  |
| No | 3633 | 71.8 |
| Yes | 1426 | 28.2 |
| **Sexual IPV** |  |  |
| No | 4521 | 89.4 |
| Yes | 538 | 10.6 |
| **Economic IPV** |  |  |
| No | 4518 | 89.3 |
| Yes | 541 | 10.7 |

In terms of lifestyle behaviors, 0.3% of the participating women consumed alcohol, and 1.0% used tobacco, however, 30.6% of them had husbands/partners who consumed alcohol. The majority of the participants were recently sexually inactive (51.7%), with one permanent sex partner in the last 12 months (95.2%), but had started sex at the age of 15-24 (82.7%) and ≤ 5-lifetime sexual partners (90.7%). Many of the women had two children (≤ 2, 64.0%), and desired to have more children (57.8%). A few women had STI symptoms in the form of a genital discharge (4.4%) and discharge (8.1%), and the majority were HIV-negative (97.7%). Most participating women had good knowledge of HIV (96.0%), no stigmatizing attitudes (77.4%), and believed that women were justified to ask their husbands to use condoms if they had STIs (85.0%). Few participants were afraid of their husbands (28.5%) and justified that other women/wives should be beaten for numerous reasons (32.1%). The majority of women had experienced IPV from their husbands/ sex partners, mostly in the form of controlling (61.2%), emotional (29.2%), physical (28.2%), economic (10.7%), and sexual violence (5.0%).

### Awareness and acceptability of PrEP among women of reproductive age in Kenya

In this study, 48.4% (95%CI: 47.2–49.7) of the women were aware of PrEP as a method of HIV prevention (**Table 2**). Additionally, a large proportion of the women approved the uptake of PrEP to prevent HIV (75% (95%CI: 73.3-76.6).

**Table 2.** Awareness and acceptability of PrEP among women in Kenya.

| **Variable** | **N** | **Weighed (95%CI)** |
| --- | --- | --- |
| PrEP awareness | 16638 | 48.4 (47.2–49.7) |
| PrEP acceptability | 7607 | 75 (73.3-76.6) |

### Factors associated with awareness of PrEP among women of reproductive age in Kenya

In this study, we found that education, ethnicity, working status, HIV status, number of sex partners, justified beating, religion, region, health decision-making, and HIV knowledge were associated with awareness of PrEP among women in Kenya (**Table 3**). The odds of participating women being aware of PrEP were 1.25 (95%CI:1.01-1.55) and 2.07 (95%CI: 1.48-2.88) times higher for those who completed secondary, and tertiary education, respectively, compared with participants who had completed primary or no education. Additionally, compared to those with poor knowledge, women who had good knowledge of HIV were 2.70 (95%CI: 1.68-4.33), times more likely to be aware of PrEP.

**Table 3.** Factors associated with awareness of PrEP among women of reproductive age in Kenya.

| Variable | PrEP awareness (N=16638) |  |  |  |
| --- | --- | --- | --- | --- |
|  | n | Yes % | uORs (95CI) | aORs(95CI) |
| **Sociodemographic factors** |  |  |  |  |
| **Age (years)** |  |  |  |  |
| 15-24 | 6140 | 15.5 | 1 | 1 |
| 25-34 | 5269 | 17.8 | **1.77 (1.58-1.99) \*\*** | 1.13 (0.91-1.41) |
| 35-59 | 5230 | 15.2 | **1.30 (1.18-1.43) \*\*** | 0.00(0.00-0.00) |
| **Residence** |  |  |  |  |
| Rural | 9801 | 24.5 | 1 | 1 |
| Urban | 6837 | 23.9 | **1.96 (1.76-2.18) \*\*** | 1.11 (0.84-1.45) |
| **Region/Province** |  |  |  |  |
| Coast | 1491 | 3.5 | 1 | 1 |
| Northeastern | 359 | 0.4 | **0.35 (0.23-0.53) \*\*** | **3.18 (1.15-8.81)\*** |
| Eastern | 2199 | 5.9 | **1.23 (1.03-1.46) \*\*** | 1.30 (0.85-1.99) |
| Central | 2180 | 6.7 | **1.63 (1.34-1.98) \*\*** | 1.47 (0.83-2.61) |
| Rift Valley | 4504 | 11.3 | 1.10 (0.95-1.27) | 1.24 (0.82-1.87) |
| Western | 1755 | 4.3 | 1.04 (0.87-1.25) | 1.31 (0.78-2.21) |
| Nyanza | 1993 | 8.0 | **3.11 (2.62-3.68) \*\*** | **2.62 (1.63-4.19)\*** |
| Nairobi | 2157 | 8.3 | **2.77 (2.11-3.63) \*\*** | 1.49 (0.86-2.56) |
| **Ethnicity** |  |  |  |  |
| Embu | 194 | 0.6 | 0.85 (0.61-1.20) | 1.00 (0.51-1.96) |
| Kalenjin | 2820 | 6.6 | **0.54 (0.47-0.64) \*\*** | 0.98 (0.67-1.44) |
| Kamba | 1913 | 6.2 | 0.99 (0.82-1.20) | 1.15 (0.64-2.06) |
| Kikuyu | 2990 | 9.7 | 1 | 1 |
| Kisii | 1001 | 3.1 | 0.91 (0.74-1.12) | 0.96 (0.60-1.52) |
| Luhya | 2711 | 7.5 | **0.73 (0.62-0.87) \*\*** | 1.09 (0.72-1.64) |
| Luo | 2187 | 9.3 | **2.10 (1.77-2.48) \*\*** | **2.58 (1.62-4.12)\*\*** |
| Maasai | 383 | 0.7 | **0.34 (0.24-0.48) \*\*** | 0.66 (0.37-1.17) |
| Meru | 987 | 2.1 | **0.47 (0.37-0.59) \*\*** | 0.77 (0.46-1.28) |
| Mijikenda/Swahili | 848 | 1.8 | **0.45 (0.36-0.57) \*\*** | 1.19 (0.66-2.13) |
| Somali | 453 | 0.6 | **0.26 (0.18-0.38) \*\*** | 0.60 (0.19-1.84) |
| Taita/Taveta | 122 | 0.3 | **0.69 (0.48-0.99) \*\*** | 0.85 (0.34-2.16) |
| Other | 6 | 0.1 | 0.67 (0.40-1.12) | **3.12 (1.23-7.92) \*\*** |
| **Education** |  |  |  |  |
| None/primary | 6967 | 15.8 | 1 | 1 |
| Secondary | 6464 | 19.0 | **1.58 (1.44-1.74) \*\*** | **1.25 (1.01-1.55) \*\*** |
| Tertiary | 3207 | 13.6 | **3.97 (3.44-4.59) \*\*** | **2.07 (1.48-2.88) \*\*** |
| **Religion** |  |  |  |  |
| Christians | 14788 | 44.1 | 1 | 1 |
| Muslims | 1183 | 2.3 | **0.49 (0.42-0.58) \*\*** | **0.51 (0.31-0.83) \*** |
| Others | 667 | 2.0 | 1.01 (0.79-1.29) | 1.13 (0.74-1.71) |
| **Marital status** |  |  |  |  |
| Unmarried (single divorced/widowed/separated) | 7354 | 20.6 | 1 | 1 |
| Married/Cohabiting | 9284 | 27.8 | **1.14 (1.04-1.25) \*\*** | 0.00(0.00-0.00) |
| **Working status/occupation** |  |  |  |  |
| Not working | 7851 | 19.7 | 1 | 1 |
| Working | 8788 | 28.8 | **1.67 (1.54-1.81) \*\*** | **1.30 (1.08-1.57)\*\*** |
| **Wealth index** |  |  |  |  |
| Poorest | 2568 | 4.6 | 1 | 1 |
| Poorer | 2956 | 7.2 | **1.60 (1.38-1.85) \*\*** | 0.97 (0.74-1.25) |
| Middle | 3069 | 8.4 | **1.96 (1.71-2.26) \*\*** | 0.97 (0.71-1.32) |
| Richer | 3722 | 11.7 | **2.60 (2.24-3.01) \*\*** | 1.38 (0.97-1.98) |
| Richest | 4323 | 16.6 | **4.16 (3.51-4.93) \*\*** | 1.55 (0.98-2.47) |
| **Sex of Household head** |  |  |  |  |
| Male | 10114 | 29.1 | 1 | - |
| Female | 6524 | 19.4 | 1.07 (0.98-1.17) | - |
| **Household number** |  |  |  |  |
| ≤4 | 7572 | 24.3 | 1 | 1 |
| ≥5 | 9066 | 24.1 | **0.69 (0.62-0.76) \*\*** | 1.03 (0.83-1.28) |
| **Health status** |  |  |  |  |
| Bad | 425 | 1.3 | 1 | - |
| Moderate | 3234 | 9.3 | 0.86 (0.65-1.15) | - |
| Good | 12979 | 37.8 | 0.88 (0.66-1.15) | - |
| **Health-seeking decision-making.** |  |  |  |  |
| Self | 3994 | 21.5 | 1 | 1 |
| Joint | 4073 | 23.4 | **1.14 (1.01-1.29) \*\*** | **1.28 (1.03-1.58)\*** |
| Partner | 1197 | 4.9 | **0.61 (0.51-0.73) \*\*** | 0.87 (0.67-1.14) |
| Others | 20 | 0.1 | **0.30 (0.10-0.88)\*\*** | 0.29 (0.05-1.65) |
| **Purchases decision making** |  |  |  |  |
| Self | 1867 | 9.9 | 1 | 1 |
| Joint | 5499 | 31.1 | 1.15 (0.99-1.33) | 0.82 (0.65-1.05) |
| Partner | 1882 | 8.8 | **0.79 (0.66-0.94) \*\*** | 0.91 (0.70-1.20) |
| Others | 37 | 0.1 | 0.56 (0.25-1.21) | 0.39 (0.12-1.24) |
| **Earnings decision making** |  |  |  |  |
| Self | 634 | 4.0 | 1 | - |
| Joint | 4510 | 27.0 | 1.17 (0.92-1.49) | - |
| Partner | 3660 | 19.5 | 0.92 (0.72-1.17) | - |
| Other | 4 | 0.014 | 0.45 (0.04-5.18) | - |
| **Husband Education** |  |  |  |  |
| None/primary | 4027 | 17.1 | 1 | 1 |
| Secondary | 2984 | 17.4 | **1.82 (1.60-2.06) \*\*** | 1.15 (0.93-1.43) |
| Tertiary | 2180 | 15.3 | **2.87 (2.38-3.45) \*\*** | 1.25 (0.94-1.66) |
| **Husband's Working status/occupation** |  |  |  |  |
| Not working | 708 | 2.6 | 1 | 1 |
| Working | 8282 | 47.2 | **2.08 (1.71-2.53)\*\*** | 1.06 (0.77-1.46) |
| **Newspaper** |  |  |  |  |
| No | 13378 | 37.0 | 1 | 1 |
| Yes | 3260 | 11.4 | **1.63 (1.45-1.84) \*\*** | 1.12 (0.90-1.38) |
| **TV** |  |  |  |  |
| No | 5613 | 12.5 | 1 | 1 |
| Yes | 11025 | 35.9 | **2.02 (1.84-2.21) \*\*** | 1.03 (0.84-1.26) |
| **Radio** |  |  |  |  |
| No | 4052 | 9.9 | 1 | 1 |
| Yes | 12586 | 38.6 | **1.53 (1.38-1.69) \*\*** | 1.21 (0.97-1.51) |
| **Internet use** |  |  |  |  |
| No | 8715 | 19.6 | 1 | 1 |
| Yes | 7923 | 28.9 | **2.59 (2.34-2.86) \*\*** | 1.03 (0.82-1.30) |
| **Lifestyle Behavioural factors** |  |  |  |  |
| **Tobacco use** |  |  |  |  |
| No | 16467 | 48.0 | 1 | - |
| Yes | 171 | 0.5 | 0.92 (0.61-1.41) | - |
| **Alcohol use** |  |  |  |  |
| No | 15733 | 47.2 | 1 | - |
| Yes | 43 | 0.1 | 1.07 (0.42-2.71) | - |
| **Husband drinks Alcohol** |  |  |  |  |
| No | 3511 | 34.0 | 1 | - |
| Yes | 1548 | 16.3 | 1.18 (0.98-1.41) | - |
| **Sexual Behavioural factors** |  |  |  |  |
| **Age at first sex (years)** |  |  |  |  |
| ≤14 | 2063 | 6.4 | 1 | 1 |
| 15-24 | 11658 | 43.7 | **1.44 (1.27-1.62) \*\*** | 1.09 (0.86-1.39) |
| ≥25 | 370 | 1.3 | 1.28 (0.93-1.77) | 0.74 (0.35-1.55) |
| **Sexual activity** |  |  |  |  |
| Inactive | 8606 | 23.3 | 1 | 1 |
| Active | 8031 | 25.1 | **1.32 (1.22-1.44) \*\*** | 1.09 (0.88-1.35) |
| **Number of sex partners in last 12 months** |  |  |  |  |
| 1 partner | 11642 | 48.8 | 1 | 1 |
| ≥2 partners | 583 | 2.9 | **1.51 (1.18-1.93) \*\*** | **2.34 (1.47-3.73)\*\*** |
| **Number of lifetime sex partners** |  |  |  |  |
| <5 | 12780 | 45.6 | 1 | 1 |
| 5-10 | 1025 | 4.4 | **1.56 (1.25-1.93) \*\*** | 0.95 (0.67-1.34) |
| >10 | 290 | 1.4 | **2.05 (1.33-3.16) \*\*** | 1.14 (0.25-5.21) |
| **HIV status** |  |  |  |  |
| Negative | 13833 | 50.7 | 1 | 1 |
| Positive | 332 | 1.7 | **2.46 (1.83-3.31) \*\*** | **4.28 (1.78-10.30)\*\*** |
| **Genital ulcer in last 12 months** |  |  |  |  |
| No | 15884 | 46.1 | 1 | 1 |
| Yes | 738 | 2.3 | 1.14 (0.93-1.38) |  |
| **Genital Discharge in the last 12 months** |  |  |  |  |
| No | 15267 | 44.1 | 1 | 1 |
| Yes | 1347 | 4.3 | **1.21 (1.03-1.43) \*** | 0.89 (0.65-1.21) |
| **Number of living children** |  |  |  |  |
| ≤2 | 10652 | 32.4 | 1 | 1 |
| 3-4 | 3916 | 11.4 | 0.92 (0.84-1.02) | 0.78 (0.60-1.01) |
| ≥5 | 2071 | 4.6 | **0.58 (0.51-0.66) \*\*** | 0.77 (0.51-1.17) |
| **Use of other HIV services** |  |  |  |  |
| **Condom use at Last Sex** |  |  |  |  |
| No | 12687 | 45.5 | 1 | 1 |
| Yes | 1408 | 6.0 | **1.45 (1.24-1.68) \*\*** | 0.95(0.57-1.59) |
| **Ever tested for HIV** |  |  |  |  |
| No | 2355 | 3.4 | 1 | 1 |
| Yes | 14283 | 45.0 | **3.45 (2.96-4.01) \*\*** | 1.46 (0.21-10.38) |
| **Pregnant** |  |  |  |  |
| No or unsure | 15712 | 45.7 | 1 | - |
| Yes I am | 926 | 2.7 | 1.03 (0.86-1.23) | - |
| **Psychosocial factors** |  |  |  |  |
| **Desire for children** |  |  |  |  |
| Wants | 9326 | 28.3 | 1 | 1 |
| Does not want | 5789 | 17.9 | 1.03 (0.95-1.12) | 1.06 (0.86-1.30) |
| Undecided | 1029 | 2.3 | **0.60 (0.51-0.71)\*\*** | 0.67 (0.44-1.02) |
| **HIV knowledge** |  |  |  |  |
| Poor | 459 | 0.6 | 1 | 1 |
| Good | 10942 | 47.9 | **5.44 (3.96-7.48) \*\*** | **2.70 (1.68- 4.33)\*\*** |
| **Would buy vegetables from vendors with HIV** |  |  |  |  |
| Yes (No stigma) | 3682 | 7.8 | 1 | 1 |
| No (stigma) | 12639 | 40.0 | **0.49 (0.44-0.55)\*\*** | 0.89 (0.71-1.11) |
| **Wife justified in asking her husband to use a condom** |  |  |  |  |
| No | 2439 | 5.4 | 1 | 1 |
| Yes | 13870 | 42.9 | **1.83 (1.63-2.07) \*\*** | 1.14 (0.91-1.44) |
| **IPV factors** |  |  |  |  |
| **Respondent afraid of Husband** |  |  |  |  |
| No | 3615 | 36.8 | 1 | - |
| Yes | 1444 | 13.5 | 0.85 (0.71-1.00) | - |
| **Justified beating** |  |  |  |  |
| No | 11282 | 36.3 | 1 | 1 |
| Yes | 5324 | 12.1 | **0.53 (0.48-0.59)\*\*** | **0.63 (0.52- 0.76)\*** |
| **IPV forms** |  |  |  |  |
| **Emotional IPV** |  |  |  |  |
| No | 3582 | 35.7 | 1 | - |
| Yes | 1477 | 14.6 | 0.99 (0.84-1.17) | - |
| **Controlling IPV** |  |  |  |  |
| No | 1963 | 19.0 | 1 | - |
| Yes | 3096 | 31.2 | 1.08 (0.93-1.26) | - |
| **Physical IPV** |  |  |  |  |
| No | 3633 | 36.2 | 1 | - |
| Yes | 1426 | 14.1 | 0.99 (0.84-1.16) | - |
| **Sexual IPV** |  |  |  |  |
| No | 4521 | 45.0 | 1 | - |
| Yes | 538 | 5.2 | 0.95 (0.73-1.25) | - |
| **Economic IPV** |  |  |  |  |
| No | 4518 | 44.9 | 1 | - |
| Yes | 541 | 5.4 | 1.02 (0.78-1.34) | - |

IPV=intimate partner violence, uOR=unadjusted odd ratios, aOR=adjusted odds ratio\* =
significant at $p < 0.05$ , \*\* = significant at $p < 0.001$ , – = not evaluated in that model, n=row
total, %= percentage of the total sample size when all the row totals are added up.

When compared with participants from the Kikuyu tribe, women from the Luo and other tribes of Kenya were 2.58 (95%CI: 1.62-4.12), and 3.12 (95%CI: 1.23-7.92) times more likely to be aware of PrEP, respectively. Similarly, participants from the Northeastern and Nyanza provinces of Kenya were 3.18 (95%CI: 1.15-8.81) and 2.62 (95%CI: 1.63-4.119) times more likely to be aware of PrEP, respectively, when compared with those from the coastal region. In addition, women who were working compared with those who were not, were more likely to be aware of PrEP (adjusted odds ratio (aOR)= 1.30 (95%CI: 1.08-1.57). Participating women living with HIV were 4.28 (95%CI: 1.78-10.30) times more likely to be aware of PrEP, compared with those without HIV infection. Similarly, women who had multiple concurrent sex partners (≥2) compared with those who had one sexual partner were 2.34 (95%CI: 1.47-3.73) times more likely to be aware of PrEP. Participating women who made health decisions jointly with their partners were 1.28 (95%CI: 1.03-1.58) times more likely to be aware of PrEP when compared to those who made decisions independently.

On the other hand, participating women who perceived that men are justified to beat their wives for numerous reasons were 0.63 (95%CI: 0.52-0.76) times less likely to be aware of PrEP than those who thought to the contrary. Likewise, women subscribing to Islam compared with the Christians were less likely to be aware of PrEP (aOR 0.51 (95%CI: 0.31-0.83).

### Factors associated with Acceptability of PrEP among women of reproductive age in Kenya

We found that age at first sex, region, and HIV knowledge were associated with the acceptability of PrEP use among reproductive-age women in Kenya (**Table 4**). Women who had their first sex at an older age (≥25 years), compared with those who had their first sex at a younger age (15-24 years), were more likely to approve the use of PrEP in the prevention of HIV (aOR 40.06 (95%CI: 14.59-109.99). Similarly, women from Nyanza province were more likely to approve the use of PrEP in the prevention of HIV, compared with their counterparts from the coastal province (aOR 8.76 (95%CI: 3.27-23.46). Lastly, compared to those with poor knowledge, women with good knowledge of HIV were 4.88 (95%CI: 1.28-18.62), times more likely to approve the use of PrEP in the prevention of HIV.

**Table 4.**
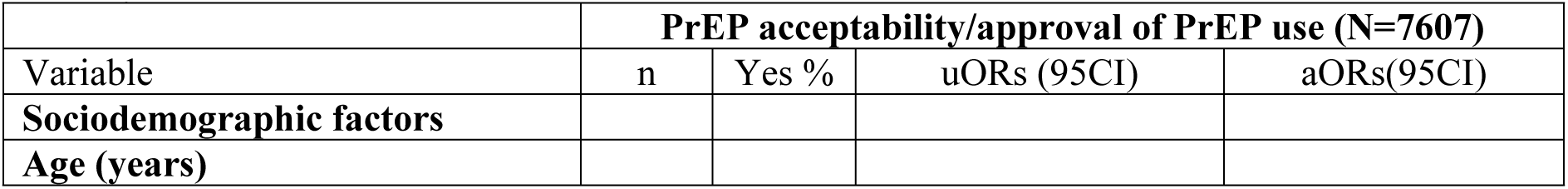
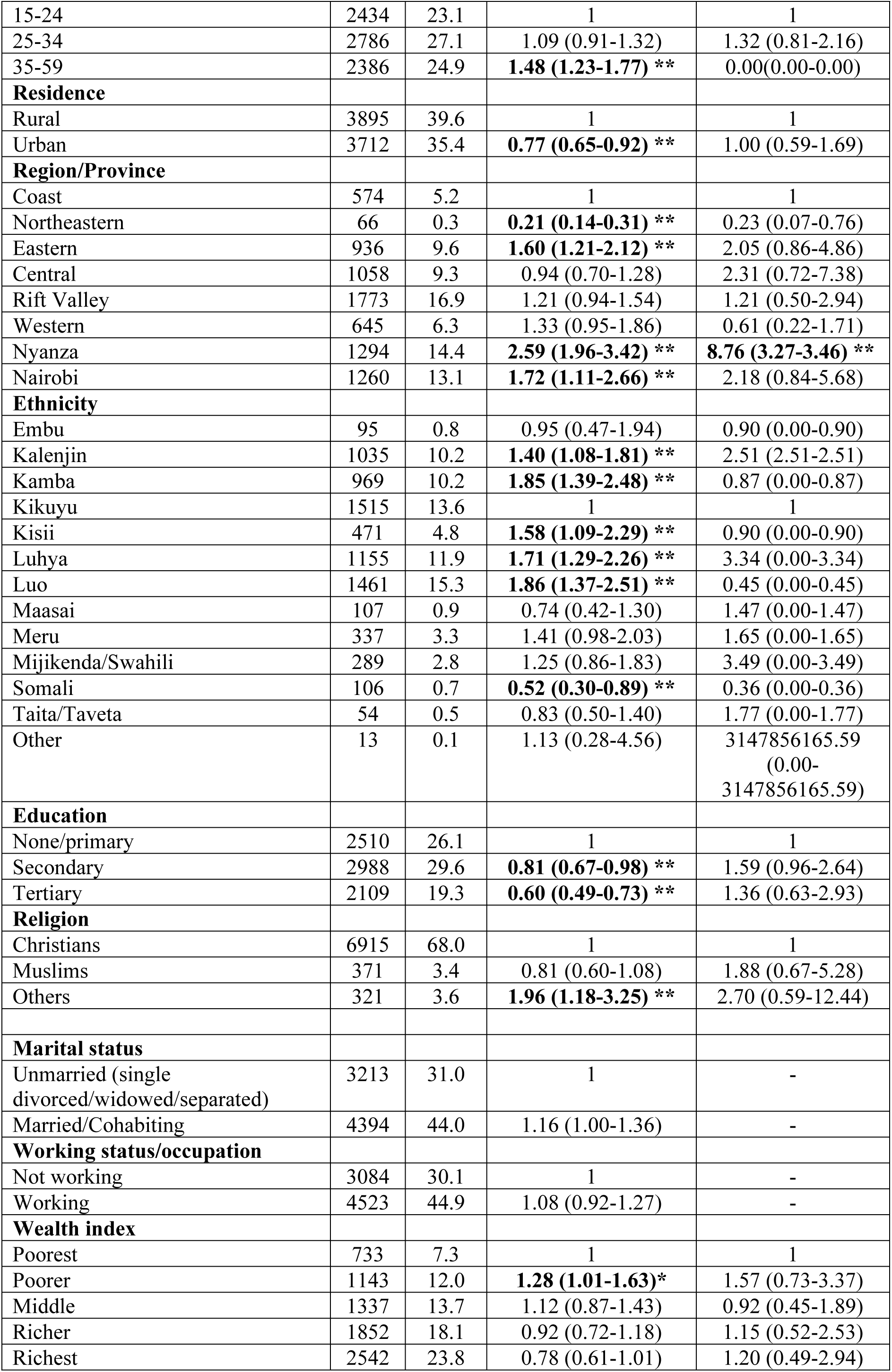
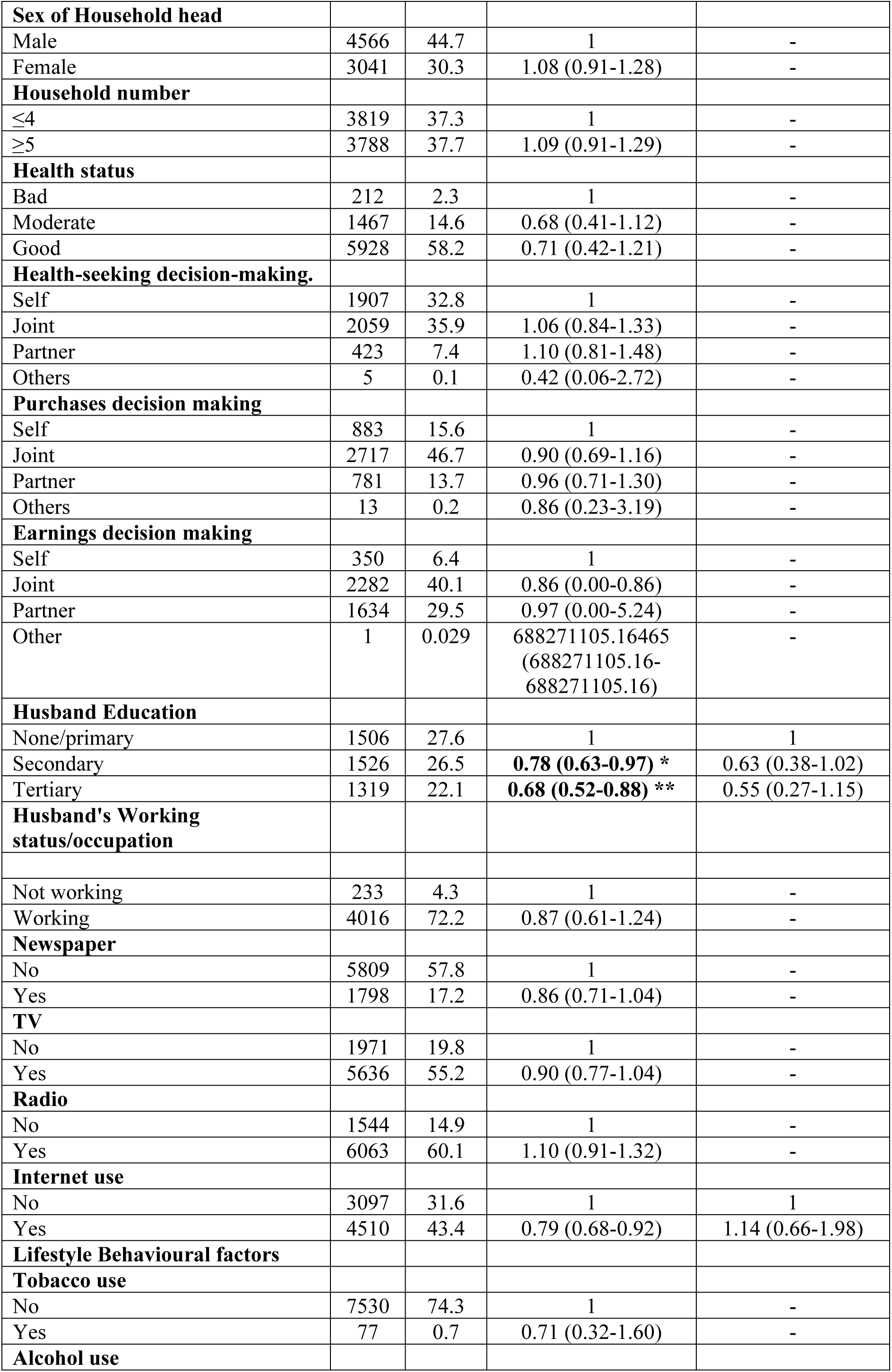
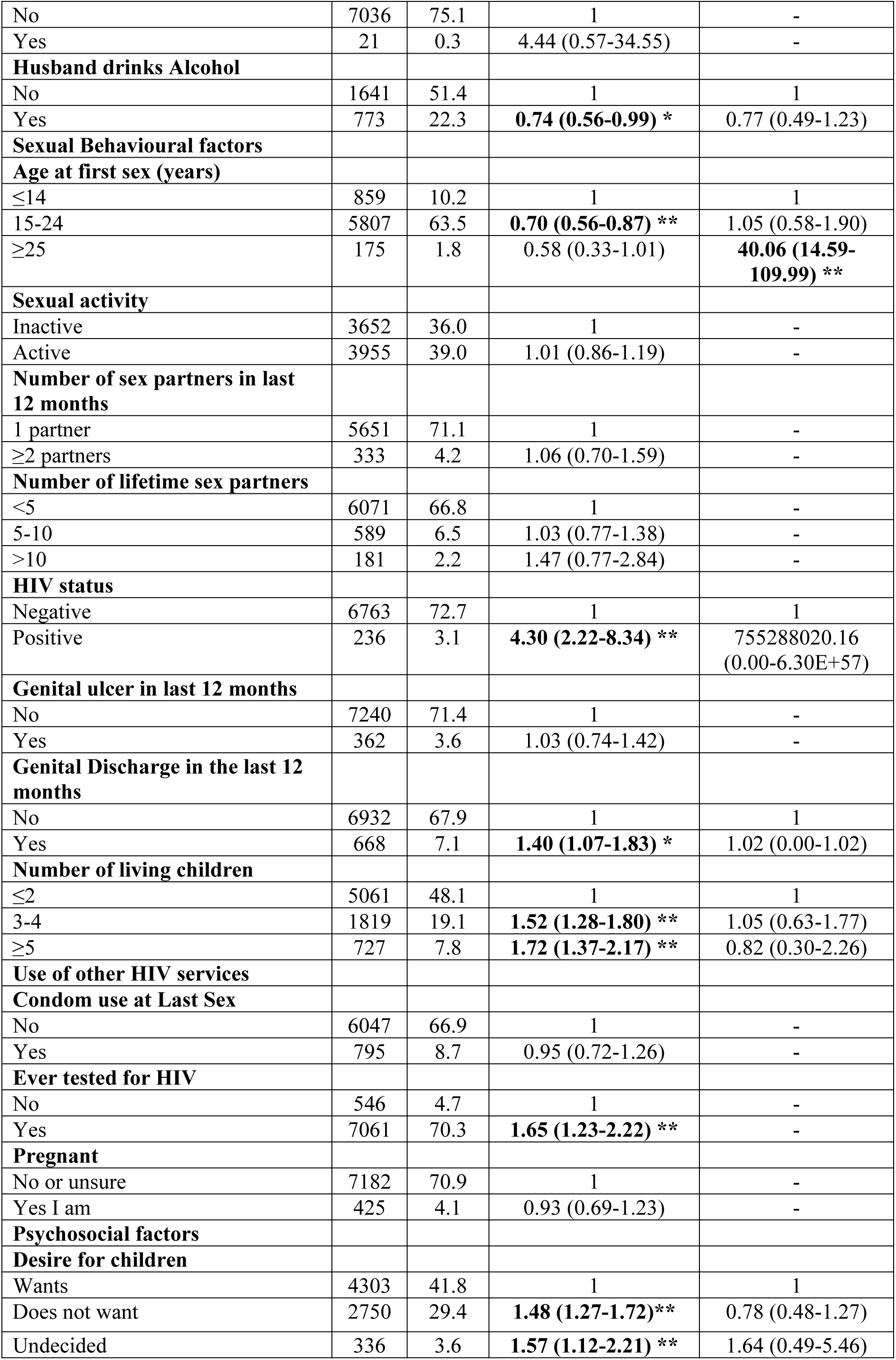
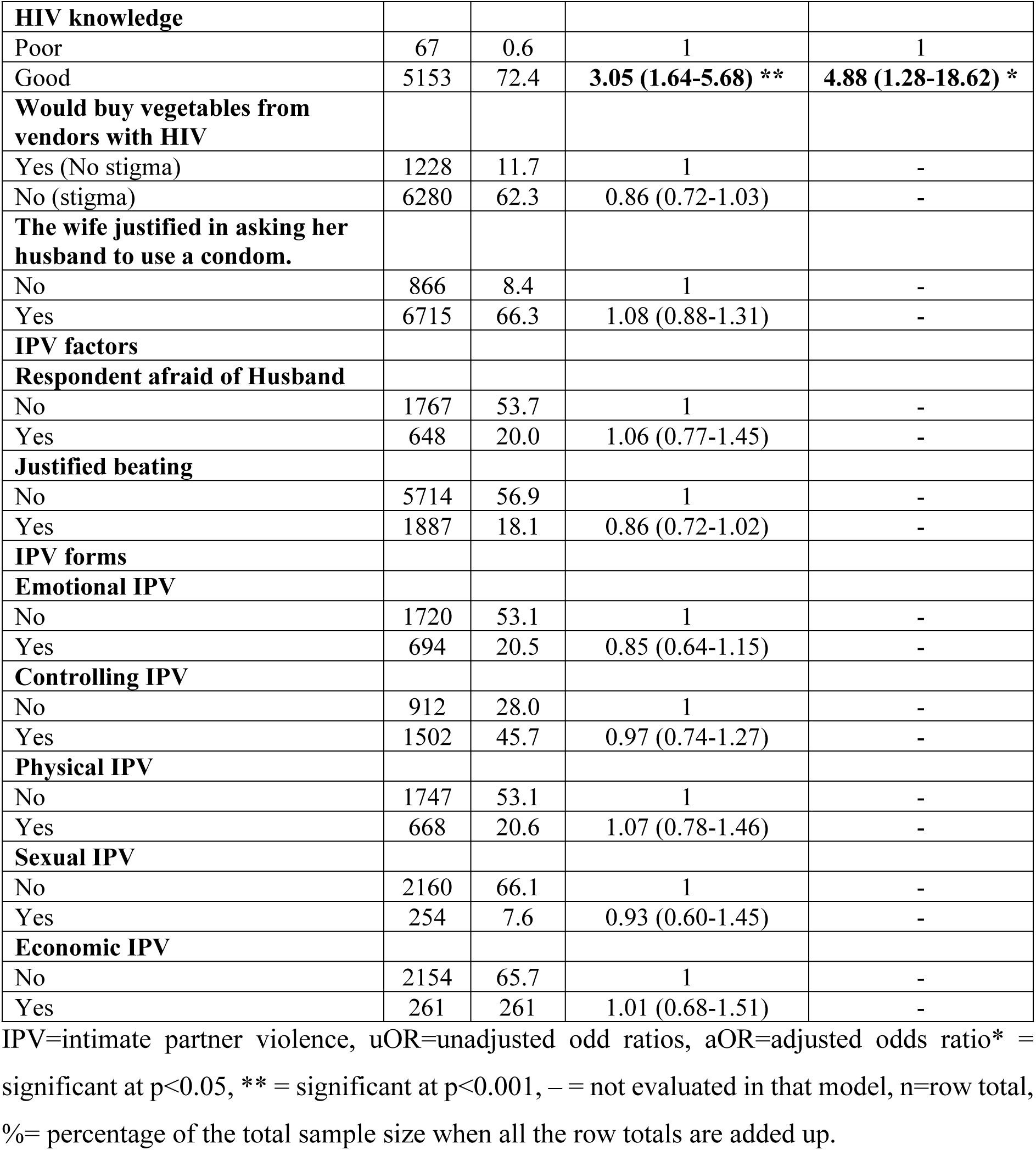
Factors associated with acceptability of PrEP among women of reproductive age in Kenya.

## Discussion

The primary objective of this study was to determine factors associated with awareness and acceptability of PrEP among WRA using an analysis of the 2022 KDHS. This study revealed that almost half (48.4%) of the women were aware of HIV PrEP, a level higher than the 13.9% reported across five SSA countries (6) and 25% observed among AGYW in sub-Saharan Africa (1). However, our observed awareness prevalence is lower than the 61.4% reported in the general AGYW population in China (18). The difference in PrEP awareness levels could be attributed to differences in the population characteristics, study methods used, variations in HIV prevention programs, public health campaigns, healthcare access, and socioeconomic factors (19, 20).

The study also found that 3 in 4 (75%) women approved the uptake of PrEP to prevent HIV. This substantial willingness to use HIV PrEP is higher than that reported among Chinese trans-women at 49.1% (18) but lower than 98% willingness among female sex workers in a study in Tanzania (21) and 91% in Nigeria by (22). This could be attributed to the characteristics of the participants but also to other factors such as risk perception, targeted education campaigns and outreaches, and access which may influence PrEP acceptability across different groups (23, 24). Therefore, we recommend that the Kenyan government, policymakers, and implementers strengthen targeted education campaigns, enhance outreach initiatives, and improve access to PrEP services while addressing risk perception to boost PrEP awareness and acceptability across diverse groups.

This study revealed significant regional disparities in awareness and acceptability of PrEP for HIV prevention, with Northeastern and Nyanza provinces showing high odds of awareness and acceptability compared with the Coastal province. This concurs with prior research highlighting the impact of geographic and sociocultural factors on HIV prevention knowledge (25). Northeastern and Nyanza’s higher acceptability and awareness is likely due to sustained HIV prevention campaigns and community health programs driven by their historically high HIV prevalence rates (26). In contrast, lower awareness in Coastal Kenya may stem from sociocultural factors, particularly among the region’s large Muslim population, where Islamic teachings emphasizing abstinence and fidelity downplay the use of biomedical interventions like PrEP, which may reduce PrEP acceptability and awareness (27, 28). These findings highlight the need for tailored, region-specific health education and interventions to address cultural and religious barriers and enhance PrEP awareness and acceptability (and subsequent PrEP uptake).

This study found that women with good HIV knowledge were more likely to be aware of and approve PrEP uptake for HIV prevention compared to those with poor knowledge, in agreement with research linking HIV literacy to increased PrEP uptake (29). Comprehensive HIV understanding boosts health-seeking behaviors, reduces stigma, and increases trust in biomedical interventions like PrEP (30, 31). Women who understand HIV transmission and prevention are more likely to see PrEP’s benefits (30, 31). Conversely, misinformation, stigma, and lack of HIV education, lower PrEP awareness and approval (32). The findings emphasize the need for expanded HIV education to improve PrEP acceptance. Strategies like community-driven campaigns, integrating PrEP into HIV prevention programs, and peer-led education could enhance women’s knowledge and approval of PrEP leading to informed decision-making about PrEP uptake.

In this study, women with secondary or tertiary education had significantly higher odds of being aware of HIV PrEP compared with those with primary or no education. These findings concur with research indicating that higher education enhances access to health information, health literacy, and engagement with healthcare services (10, 18). Limited education may restrict exposure to HIV prevention resources, perpetuating awareness gaps (33). The Kenyan government and stakeholders should prioritize promoting women’s education to address this issue. Targeted community outreaches and policy reforms are essential to enhance educational access and quality (34). Advancing gender equality in education can further empower women to engage with health services boosting PrEP awareness, acceptability, and subsequent uptake.

Women who were working were more likely to be aware of HIV PrEP compared to those not working, consistent with research linking socioeconomic factors like employment to increased awareness of HIV prevention strategies (35, 36). Employment provides greater access to health information, healthcare services, and workplace wellness programs, which enhance PrEP awareness (37). Unemployed women, however, face economic and educational barriers that restrict their exposure to HIV prevention resources (38). These findings emphasize the need for socioeconomic empowerment interventions for women to improve PrEP awareness. Workplace health education programs can also promote PrEP awareness among employed women. Targeted outreach efforts are crucial for unemployed women to overcome PrEP awareness barriers bridging disparities in HIV prevention information access.

When compared with participants from the Kikuyu tribe, women from the Luo and other tribes of Kenya were 2.51, and 3.27 times more likely to be aware of PrEP, respectively. This finding concurs with research that revealed that tribal characteristics, including cultural beliefs, social norms, and access to resources, significantly influence HIV prevention program awareness (6). This study’s finding related to ethnic differences in PrEP awareness may be attributed to the lower HIV prevalence among the Kikuyu ethnic group compared to other tribes. For example, the Kikuyu who predominantly occupy central Kenya whose HIV prevalence stands at 4.8% compared with the Luo ethnic group who predominantly occupy Nyanza province with a prevalence of HIV at 13.6% (26). Therefore, this study’s findings highlight the need for culturally sensitive PrEP awareness campaigns, and improved access, ensuring inclusive HIV prevention strategies across diverse ethnic groups in Kenya.

Relatedly, participating women living with HIV were 3.79 times more likely to be aware of PrEP, compared with those without HIV infection. There is not much existing information about whether women living with HIV are more likely to be aware of PrEP than women without HIV. However, this heightened awareness can be attributed to their frequent engagement with healthcare services, where PrEP education is integrated into routine HIV care and prevention counseling (Kumah et al., 2023). Additionally, targeted interventions, including peer support networks and HIV prevention programs, further contribute to their knowledge of PrEP (39). In contrast, HIV-negative women may have limited exposure to the healthcare system and to PrEP education, underscoring the need for expanded community outreach and awareness campaigns to bridge this gap. More qualitative research is needed to explore the disparity to inform strategies to enhance PrEP awareness among HIV-negative women.

Women with multiple concurrent sexual partners (≥2) were 2.29 times more likely to be aware of PrEP compared with those with a single-sex partner. These findings concur with a study conducted among WRA in the USA, which reported a similar association between multiple sexual partnerships and increased PrEP awareness (38). This suggests that women engaging in high-risk sexual behaviors may have greater exposure to HIV prevention messaging or healthcare services promoting PrEP (38). Also, such risk groups may have a heightened sense of perceived HIV risk hence seeking preventive measures (40). To bridge gaps in awareness and uptake, inclusive educational campaigns and community engagement targeting the general population should be implemented to improve awareness of HIV preventive services like PrEP to the whole population regardless of the degree of HIV exposure or risk.

Participating women who perceived that men are justified to beat their wives for numerous reasons were 0.63 (95%CI: 0.52-0.76) times less likely to be aware of PrEP than those who thought to the contrary. This study revealed that women who perceived that men are justified to beat their wives for numerous reasons were less likely to be aware of PrEP compared with those who reject such beliefs, consistent with research linking patriarchal norms to reduced autonomy in health-related decision-making, including access to HIV prevention services (41, 42). These women often face limited exposure to HIV prevention messaging due to lower healthcare-seeking behaviors, restricted decision-making, and fear of partner repercussions. In settings where IPV is normalized, healthcare providers may also be less likely to discuss PrEP, further reducing awareness (43). Gender-transformative interventions are essential to address these barriers. Community education, male engagement initiatives, and policies tackling gender-based violence should be integrated into HIV prevention efforts. Incorporating IPV screening and counseling into PrEP delivery can support at-risk women, enhancing awareness and uptake.

This study found that Muslim women in Kenya are less likely to be aware of PrEP for HIV prevention compared with Christian women, in agreement with research on sociocultural and religious influences on health behaviors, particularly in relation to HIV prevention (27, 44). Limited autonomy, male guardianship, and conservative norms limiting sexual health discussions contribute to lower PrEP awareness among Muslim women (27). Perceptions of reduced HIV risk due to male circumcision and emphasis on abstinence further discourage PrEP engagement in Muslim communities (44). Culturally sensitive interventions are needed to address these barriers (45). Engaging minority faith-based leaders and offering non-judgmental PrEP counseling can boost awareness about PrEP (46). Leveraging discreet digital health campaigns and community-based education tailored to religious contexts can enhance PrEP awareness.

Additionally, women who made health decisions jointly with their partners were 1.28 times more likely to be aware of HIV PrEP compared with those who made them independently, in agreement with findings on research on the role of dyadic-level influence and relational autonomy in HIV prevention (47, 48). Joint decision-making often indicates healthier and unrestrained relationships that enhance access to HIV prevention information. In contrast, independent decision-making, possibly due to limited partner support or power imbalances, may restrict such access (49). These findings suggest the need for strategies that promote couple communication and shared responsibility. Community-based interventions and couple-centered counseling can foster relational autonomy. Integrating PrEP education into family planning or antenatal services could further boost awareness. Engaging male partners is crucial in contexts where their involvement significantly influences such health outcomes.

This study found that women who had their first sexual experience at ≥25 years were 40 times more likely to approve PrEP for HIV prevention compared with those who initiated sex at 15– 24 years, consistent with research linking early sexual debut to lower uptake of HIV prevention tools (50). Women who initiate sexual activity later may exhibit greater health awareness and autonomy, contributing to high PrEP acceptability (31). Younger women with early sexual debut may face barriers like limited PrEP awareness, stigma, and inadequate access to youth-friendly health services. These findings underscore the need for targeted interventions, such as comprehensive sex education and youth-friendly PrEP delivery models, to address the unique needs of early sexual initiators and improve HIV prevention uptake among younger populations.

## Strength and limitations

This study is based on the latest nationally representative data from the 2022 Kenya Demographic and Health Survey (KDHS), ensuring generalizability, reliability, and comparability through its large sample size and standardized data collection methods. However, its reliance on retrospective self-reported data introduces potential recall and social desirability biases. Additionally, the cross-sectional design limits the ability to establish causal relationships and capture evolving trends in PrEP awareness and acceptability among WRA. Another limitation of this study is that it focuses solely on PrEP awareness and acceptability rather than actual uptake and usage, limiting its ability to assess real utilization. Despite this limitation, the results from this study can be used to draw conclusions and predict the likely trend in PrEP utilization among WRA.

## Conclusions

While 48.4% of WRA were aware of PrEP and 75% approved its use, this is still suboptimal and was linked to disparities in education, employment, cultural norms, and regional contexts. Factors like higher education, working status, regional disparities, and good HIV knowledge significantly predicted high odds of PrEP awareness and approval. Conversely, cultural barriers, such as justified wife beating and religious affiliations, particularly Islam, predicted lower odds of PrEP awareness. Therefore, to improve PrEP awareness, acceptance, and subsequent uptake, interventions should focus on expanding education, leveraging mass media, addressing gender-based barriers, and integrating PrEP services into healthcare systems. Culturally sensitive approaches and targeted efforts in underserved regions are essential to ensure equitable access and contribute to ending HIV as a public health threat by 2030.

## Abbreviations

NASCOP: National AIDS and STI Control Programme
KDHS: Kenya Demographic and Health Survey
HIV/AIDS: Human Immunodeficiency Virus/ Acquired Immunodeficiency Syndrome
PrEP: Pre-exposure prophylaxis
UNAIDS: United Nations Programme on HIV and AIDS
WHO: World Health Organization
MOH: Ministry of Health
WRA: Women of Reproductive Age
MSM: Men Who Have Sex with Men
AGYW: Adolescent Girls and Young Women
PWID: People Who Inject Drugs
ICF: Inner-city Fund
CI: Confidence interval
aOR: Adjusted odds ratio
EA: Enumeration area

## Data Availability

In this study, the data set used is freely available upon seeking permission from the MEASURE DHS website (URL: https://www.dhsprogram.com/data/available-datasets.cfm). However, the authors are not authorized to share this dataset with the public. In case one is interested in the dataset, one can seek it with written permission from the MEASURE DHS website (URL: https://www.dhsprogram.com/data/available-datasets.cfm).

## Acknowledgments

We thank the DHS program for making the data available for this study.

## DECLARATIONS

### Authors’ contributions

J.B.A and L.N Conceived the idea, drafted the manuscript, performed analysis, interpreted the results, and drafted the subsequent versions of the manuscript. E.A, R.K, A.N, I.N and J.K reviewed the first draft, helped in results interpretation, and drafted the subsequent versions of the manuscript. All authors read and approved the final manuscript.

### Ethics approval and consent to participate

High international ethical standards are fully ensured during MEASURE DHS surveys and the study protocol is performed following the relevant guidelines. The 2022 KDHS survey protocol was reviewed and approved by the ICF Institutional Review Board. Then, written informed consent was obtained from human participants and written informed consent was also obtained from legally authorized representatives of minor participants.

### Consent for publication

This is not applicable in this study.

